# The Development and Validation of A1c+, a novel multivariable prediction model for the diagnosis of diabetes

**DOI:** 10.1101/2025.02.10.25321897

**Authors:** Alan L. Hutchison, Simar Narula, Mary E. Rinella, Brandon Faubert, Raghavendra G. Mirmira, William F. Parker

**Affiliations:** Section of Gastroenterology, Hepatology and Nutrition, University of Chicago Medicine, Chicago, IL, 60637; Section of Hematology and Oncology, University of Chicago Medicine, Chicago, IL, 60637; Section of Endocrinology, Diabetes and Metabolism, University of Chicago Medicine, Chicago, IL, 60637; Section of Pulmonary and Critical Care, Department of Medicine, University of Chicago Medicine, Chicago, IL, 60637

## Abstract

**Importance:** The hemoglobin A1c and fasting plasma glucose (FPG) have known limitations for diabetes diagnosis, but models to identify individuals who would benefit from 2-hour oral glucose tolerance testing (OGTT) are limited.

**Objective:** To determine if OGTT-only diagnosed diabetes has comparable outcomes to A1c- or FPG-diagnosed diabetes and if standard clinical features could be leveraged to identify undiagnosed diabetes.

**Design:** Multivariable prediction model development and validation.

**Setting:** US National Health and Nutrition Examination Survey (NHANES) data with corresponding US National Center for Health Statistics (NCHS) mortality data.

**Participants:** Of 105,862 NHANES subjects from 1999 to 2016, we identified 13,800 subjects with FPG, A1c, or OGTT results (11,550 with mortality data) and 92,062 other subjects (53,255 with mortality data).

**Exposure:** OGTT-diagnosed diabetes

**Main Outcomes and Measures:** The primary outcomes were association of mortality with diabetes diagnostic approach and models to diagnose diabetes. We used a gradient boosted machine decision tree to predict diabetes from standard clinical features. In the test set, we compared the AUROC and the net benefit by decision curve analysis to A1c, FPG, and a combination of the two. We performed survival analysis based on method of diabetes diagnosis and diabetes model predictions.

**Results:** The rate of OGTT-only diabetes was 1.34%. Subjects with OGTT-only diabetes had equivalent risk of mortality compared to subjects with FPG- or A1c-diagnosed diabetes after adjusting for age, sex, and race/ethnicity. A model using the A1c and standard clinical features (A1c+ model) outperformed the A1c to exclude diabetes (Sensitivity at Youden’s Index: 0.72 vs. 0.37). Adding FPG to that model (A1c/FPG+) outperformed FPG for excluding diabetes (Sensitivity: 0.87 vs. 0.48). Subjects with A1c/FPG+-predicted diabetes but sub-diagnostic A1c and FPG had equivalent mortality (HR=8.2, p<2*10^−16^) to those with A1c or FPG-diagnosed diabetes (comparison p<0.17).

**Conclusions and Relevance:** Diabetes diagnosed by OGTT alone has equivalent mortality to A1c and FPG-diagnosed diabetes. A model using standard clinical features can bolster the A1c and FPG to identify potentially undiagnosed diabetes. Model predictions associated with mortality equivalently to having a diabetes diagnosis. Implementation of a clinical decision support tool could improve diagnosis of diabetes and lead to earlier interventions.

**Key Points:** *Question:* Are individuals with diabetes only diagnosable by oral glucose tolerance test (OGTT) at similar risk of mortality as those diagnosed by hemoglobin A1c or fasting plasma glucose (FPG)? Can readily-available clinical data improve diagnosis?

*Findings:* Stratifying NHANES subjects by OGTT-only (1.34%) vs A1c or FPG diagnosis (4.13%) of diabetes found that OGTT-only diagnosis had equivalent mortality. A model combining A1c and standard clinical features (A1c+) had superior AUROC for diabetes compared to the A1c alone. The addition of FPG (A1c/FPG+) had superior AUROC compared to the FPG and A1c+. The A1c/FPG+ predictions strongly associated with mortality in subjects with sub-diagnostic A1c and FPG.

*Meaning:* Incorporation of clinical features can improve diabetes diagnosis missed by A1c and FPG. Model prediction of diabetes associates with mortality.

## Introduction

The diabetes epidemic continues to grow in the United States, impacts an estimated 38.4 million people (8.7 million likely undiagnosed), and generates health care costs of $412.9 billion per year^2,3^. The American Diabetes Association (ADA) recommends the hemoglobin A1c (A1c), fasting plasma glucose (FPG), or oral glucose tolerance test (OGTT) for the diagnosis of diabetes. The A1c has become the predominant method clinically due to its ease of use^4,5^. However, A1c is inaccurate in the setting of chronic diseases such as chronic kidney disease, heart failure, cirrhosis, and after solid organ transplant^6,7^, which are all increasing in prevalence^2,8–10^. FPG is more sensitive in some populations in which the A1c is inaccurate, but requires fasting. OGTTs are recommended in these patient populations^11^, but application at the population-level is hindered by high rates of non-adherence^12,13^. It is unclear how concerning this is for the general population: the current number of US residents in whom diabetes is only diagnoseable by OGTT (OGTT-only) is unknown, as is the current relevance of OGTT-only diabetes. Here, we investigate the rate of OGTT-only diabetes and its association with mortality.

If OGTT-only diabetes is significant – identifying patients with A1c < 6.5% and FPG < 126 mg/dL who need OGTT would be challenging, and would need to be performed by already-taxed^14^ general practitioners. Clinical decision support tools could aid in this detection^15,16^. This used to be a laborious process, but with the widespread adoption of the electronic medical record significant amounts of patient data are readily accessible within these systems, allowing for machine learning solutions to this diagnostic dilemma. A powerful machine learning approach, decision tree analysis, overcomes the problems of collinearity and missing data that impact large dataset analysis. The premiere decision tree algorithm is a gradient-boosted decision tree analysis, XG Boost^17^, which has been used to predict outcomes such as sepsis, heart failure mortality, and decompensated cirrhosis^18–20^. Prior machine learning approaches have improved detection of diabetes, but these have focused on patient-reported history ^21,22^ as their predictive label, as opposed to the clinical difficult-to-obtain 2 hour glucose from an OGTT. Liu *et al.* use the 2-hour glucose in a Chinese population, but focus on patient history questions more than standard lab values, and do not focus on chronic diseases ^23^. Here, we develop an XG Boost-based machine learning model to predict diabetes using A1c, features normally obtained during standard care, and with and without FPG, which would guide ordering of an OGTT.

In this work, we aim to understand the association of diabetes diagnosed by OGTT alone with mortality, to develop and validate machine learning multivariate prediction models to identify diabetes missed by A1c and FPG but diagnosed by OGTT, and to assess the association of predicted diabetes with mortality.

## Methods

This study used data from the National Health And Nutrition Examination Survey (NHANES), a national survey of U.S. residents. A subset of subjects in NHANES from 1999-2016 received A1c and FPG measurements and a subset from 2005-2016 received A1c, FPG, and OGTT measurements. Of these, many have corresponding mortality data available from the National Center for Health Statistics. Data were divided into the training (NHANES years 2005-2014) and evaluation (NHANES years 2015-2016) datasets, in accordance with the TRIPOD-AI guidelines^24^.

### Primary outcome: Diagnosis of diabetes

The primary outcome was the diagnosis of diabetes based on A1c greater than or equal to 6.5%, FPG greater than or equal to 126 mg/dL, and/or two-hour glucose greater than or equal to 200 mg/dL. For national estimates we resampled the NHANES data using fasting or OGTT-specific weights.

### Secondary outcome: Survival

The secondary outcome was survival based on linked mortality data from the NCHS.

### Predictor variable extraction

We extracted a set of standard clinical features, including vital signs, anthropometrics, baseline, fasting, and additional labs (Supplementary Methods, Supplementary Table 1) from NHANES.

To increase the predictive ability of our model, we excluded factors that were missing in more than 12% of the data and removed overlapping variables (Supplementary Table 2). We focused on features that would be already collected during routine standard of care for U.S. adults. In the standard of care model, we included vital signs, patient history, complete blood count and differential, comprehensive metabolic panel, and hemoglobin A1c. A complete blood count and comprehensive metabolic panel are required for screening for metabolic-dysfunction associated steatotic liver disease (MASLD, previously NAFLD), which should be done for patients at risk of pre-diabetes or diabetes^1,25^. We used OGTT-specific NHANES sample weights in our model training.

### Model Development

We used gradient-boosted decision tree analysis (XG Boost) to develop two versions of our model. A1c+ included the A1c and all non-fasting lab results and other features described above. A1c/FPG+ additionally included the fasting plasma glucose. See the Supplemental Methods for further information.

### Model evaluation

For model comparison, the model test characteristic diagnostic thresholds were determined using Youden’s index. In the evaluation dataset, we compared 1) A1c+ non fasting to standard approach of A1c ≤ 6.5% and 2) A1c+ fasting to the standard approach of A1c ≤ 6.5% or FPG ≤ 126 mg/dL. We compared models based on input characteristics, area-under-the-receiver-operating-curve, sensitivity, specificity, and net benefit.

### Survival analysis

We generated Kaplan-Meier curves to estimate and visualize survival probabilities according to mode of diagnosis. We tested differences in survival based on diagnosis and prediction of diabetes using survey-weighted Cox proportional hazard regression models to estimate hazard ratios (HRs) and 95% confidence intervals (CIs) for all-cause mortality. We performed unadjusted and age, sex, and race/ethnicity-adjusted analysis comparing mode of diabetes diagnosis. As model-based diabetes predictions incorporated age, sex, and race/ethnicity, we did only unadjusted analysis for the model-based survival analyses. To account for the weighted sample, we used masked variance pseudo-strata (PSU) and masked variable pseudo-PSU in addition to MEC sample weights, fasting sample weights, and OGTT sample rates where appropriate. See the Supplemental Methods for further information.

### Sensitivity analyses

One potential limitation of the A1c and FPG thresholds is that they may not be calibrated to a modern 2005-2016 population. To account for this and to further validate the strength of our models, we performed sensitivity analyses where we allowed the A1c and FPG to vary, thereby generating an AUROC for the A1c and FPG. Likewise, we generated an XGBoost model using the A1c and FPG (A1c+FPG) alone for comparison.

## Results

### Patient characteristics

From 1999 to 2016, 105,862 subjects had laboratory measurements, but only 13,800 of the 60,936 subjects from 2005-2016 received OGTT testing (STROBE Diagram, Supplementary Figure 1). Of those 13,8000 subjects, the mean age of the cohort was 40 years (± 20.5) and 49.8% were female. 42% subjects were non-Hispanic white, 20% Black, 18% Mexican-American, 10% other Hispanic, and 10% other / multi-racial. The average height was 167 cm (± 10), the average weight was 78 kg (± 22), the average waist circumference was 95 cm (± 17), and the average systolic and diastolic blood pressures were 120 mmHg (± 17) and 67 mmHg (± 13), respectively. The average FPG was 100.2 mg/dL (± 17.9), the average A1c was 5.4% (±0.6), and the average two-hour glucose was 117 mg/dL (± 49). Table 1 shows the summary top twenty features in the FPG/A1c+ model (discussed below) for the 2005-2014 and 2015-2016 cohorts, as well as for the subjects from 1999-2016 who did not have OGTT testing.

**Table 1:** Top 20 features of interest in A1c/FPG+ model to predict diabetes for the non-OGTT 1999-2016 cohort, OGTT 2005-2014 cohort (used as a training cohort), and the OGTT 2015-2016 (used as a validation cohort).

|  | 1999-2016,<br>No OGTT | 2005-2014,<br>OGTT | 2015-2016,<br>OGTT |
| --- | --- | --- | --- |
| N | 92,062 | 11,718 | 2,082 |
| **Fasting Plasma Glucose** |  |  |  |
| Mean (SD) | 108.1 (±43.1) | 101.8 (±18.0) | 102.6 (±18.9) |
| Missing | 77,452 (84.1%) | 0 (0%) | 0 (0%) |
| **Age** |  |  |  |
| Mean (SD) | 29.0 (±25.1) | 41.1 (±20.6) | 41.8 (±20.3) |
| Missing | 13,842 (15.0%) | 0 (0%) | 0 (0%) |
| **Hemoglobin A1c** |  |  |  |
| Mean (SD) | 5.6 (±1.1) | 5.4 (±0.6) | 5.5 (±0.6) |
| Missing | 46,967 (51.0%) | 0 (0%) | 0 (0%) |
| **Waist Circumference** |  |  |  |
| Mean (SD) | 84.6 (±22.4) | 94.4 (±16.7) | 95.9 (±17.5) |
| Missing | 28,576 (31.0%) | 189 (1.6%) | 41 (2.0%) |
| **Systolic Blood Pressure** |  |  |  |
| Mean (SD) | 119.0 (±19.6) | 119.2 (±16.8) | 121.7 (±17.5) |
| Missing | 39,054 (42.4%) | 258 (2.2%) | 26 (1.2%) |
| **Height** |  |  |  |
| Mean (SD) | 153.7 (±24.6) | 167.4 (±10.1) | 166.3 (±10.1) |
| Missing | 25,601 (27.8%) | 73 (0.6%) | 7 (0.3%) |
| **% Segmented Neutrophils** |  |  |  |
| Mean (SD) | 54.4 (±12.1) | 56.1 (±9.9) | 55.8 (±9.4) |
| Missing | 30,629 (33.3%) | 43 (0.4%) | 9 (0.4%) |
| **% Lymphocytes** |  |  |  |
| Mean (SD) | 33.9 (±11.1) | 31.9 (±8.8) | 32.2 (±8.7) |
| Missing | 30,629 (33.3%) | 43 (0.4%) | 9 (0.4%) |
| **ALT** |  |  |  |
| Mean (SD) | 24.0 (±24.0) | 24.5 (±19.8) | 23.6 (±14.0) |
| Missing | 47,828 (52.0%) | 99 (0.8%) | 6 (0.3%) |
| **Platelets** |  |  |  |
| Mean (SD) | 276.9 (±75.9) | 251.4 (±66.4) | 237.5 (±58.8) |
| Missing | 30,476 (33.1%) | 22 (0.2%) | 9 (0.4%) |
| **AST** |  |  |  |
| Mean (SD) | 25.2 (±16.9) | 25.7 (±20.0) | 24.7 (±10.8) |
| Missing | 47,830 (52.0%) | 102 (0.9%) | 6 (0.3%) |
| **Creatinine** |  |  |  |
| Mean (SD) | 0.8 (±0.4) | 0.8 (±0.3) | 0.8 (±0.3) |
| Missing | 47,748 (51.9%) | 81 (0.7%) | 6 (0.3%) |
| **Diastolic Blood Pressure** |  |  |  |
| Mean (SD) | 66.2 (±13.9) | 67.1 (±11.9) | 67.2 (±11.6) |
| Missing | 39,264 (42.6%) | 309 (2.6%) | 32 (1.5%) |
| **Poverty Ratio** |  |  |  |
| Mean (SD) | 2.2 (±1.6) | 2.5 (±1.6) | 2.4 (±1.6) |
| Missing | 20,797 (22.6%) | 817 (7.0%) | 193 (9.3%) |
| **Total Protein** |  |  |  |
| Mean (SD) | 7.3 (±0.5) | 7.2 (±0.5) | 7.2 (±0.4) |
| Missing | 47,797 (51.9%) | 89 (0.8%) | 6 (0.3%) |
| **Hemoglobin** |  |  |  |
| Mean (SD) | 13.7 (±1.5) | 14.3 (±1.5) | 14.1 (±1.5) |
| Missing | 30,475 (33.1%) | 21 (0.2%) | 9 (0.4%) |
| **Arm Circumference** |  |  |  |
| Mean (SD) | 27.5 (±8.0) | 32.1 (±5.3) | 32.3 (±5.4) |
| Missing | 22,117 (24.0%) | 142 (1.2%) | 28 (1.3%) |
| **Pulse** |  |  |  |
| Mean (SD) | 75.1 (±12.9) | 72.1 (±11.8) | 72.3 (±11.4) |
| Missing | 38,525 (41.8%) | 220 (1.9%) | 25 (1.2%) |
| **% Monocytes** |  |  |  |
| Mean (SD) | 8.0 (±2.4) | 8.1 (±2.4) | 8.2 (±2.1) |
| Missing | 30,629 (33.3%) | 43 (0.4%) | 9 (0.4%) |
| **Weight** |  |  |  |
| Mean (SD) | 58.4 (±32.2) | 77.8 (±21.5) | 78.5 (±22.0) |
| Missing | 19,227 (20.9%) | 67 (0.6%) | 8 (0.4%) |

### National prevalence of diabetes by diagnosis type

Resampling the 13,800 subjects based on NHANES-assigned sampling weights, the prevalence of diabetes diagnosed by any method was 5.46%, while 1.34% of the population had diabetes diagnosed by OGTT alone. Supplementary Table 3 shows the confusion matrix for the diagnosis of diabetes and pre-diabetes. To better understand this discordance, we compared the A1c to the two-hour glucose in individuals with an A1c less than 6.5% in the setting of different comorbidities: chronic kidney disease, anemia, obesity, liver fibrosis, age, and hypertension. The relationship between the A1c and the two-hour glucose varied widely across these (Supplementary Figure 2).

### Association of diabetes diagnosis with mortality

In the 2005-2016 cohort, 11,500/13,8000 subjects had paired NCHS mortality data. We separated subjects by no diagnosis of diabetes, diagnosis of diabetes by at least A1c or FPG, and diagnosis of diabetes by OGTT alone. We found that subjects with OGTT-only diabetes had a 5 times increased risk of mortality compared to those without diabetes (p<2×10^−16^) and a 1.3 times increased risk of mortality compared to those with diabetes diagnosed by A1c or FPG (independent of if their OGTT was also diagnostic) (p<0.03). When we adjusted for age, sex, and race/ethnicity these effect sizes decreased (OGTT-only vs no diabetes: HR=1.6, p<0.0004; OGTT-only vs A1c/FPG diabetes: HR=1.0, p=0.98) (Figure 1A). OGTT-only diabetes had equivalent odds of resulting in diabetes being a leading cause of death (Figure 1B), but lower rates of resulting in diabetes being a contributing cause of death (Figure 1C). This finding of a substantial survival difference for OGTT-only diabetes, even if accounted for by age, sex, and race/ethnicity, as well as equivalent rates of diabetes being the leading cause of death, motivated us to develop a model to detect diabetes missed by A1c or FPG that would include such demographic data.

**Figure 1.**
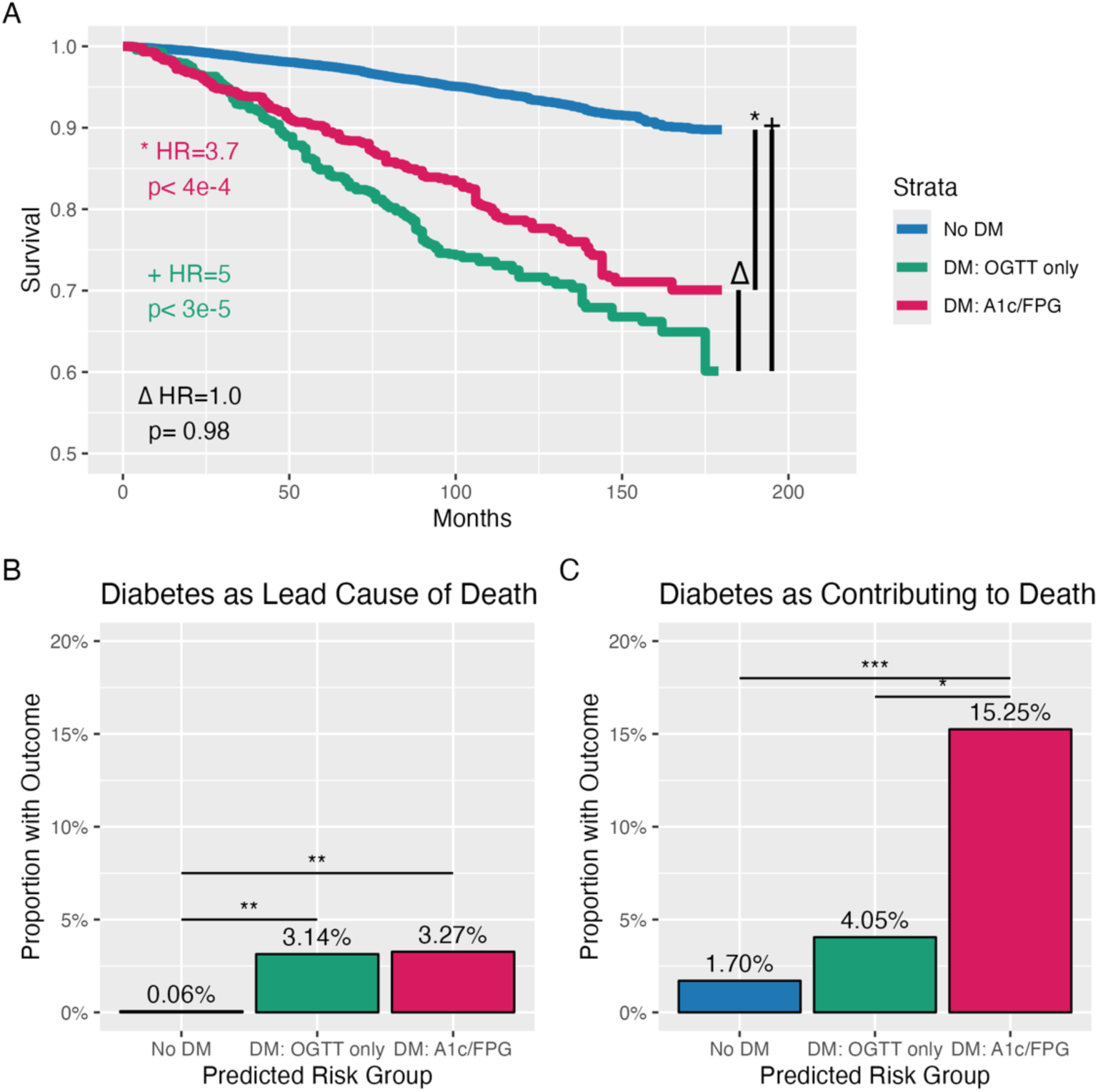
The subjects with diabetes diagnosed via OGTT alone had equivalent mortality to those with diabetes diagnosed with A1c and/or FPG (A), though they equivalent rates of diabetes being the leading cause of death (B) and they had lower rates of diabetes as a contributing cause of death, though this was still higher than those without a diagnosis of diabetes altogether (C). Comparison of mortality (A), diabetes as a leading cause of death (B), and diabetes as a contributing cause of death (C) from NCHS data for NHANES 2005-2016 cohorts that had OGTT measurement stratified by subjects with a diagnosis of diabetes by A1c ≥ 6.5% or FPG ≥ 126 mg/dL (red), subjects with a diagnosis of diabetes by OGTT alone (2-hour glucose > 200 mg/dL but A1c < 6.5% and FPG < 126 mg/dL), or subjects without a diagnosis of diabetes. (A) Kaplan-Meier curves of mortality stratified by presence and diagnostic method of diabetes. Comparisons made adjusting for age, sex, and race/ethnicity. For the * and + comparisons the reference is to subjects without diabetes. For the Δ comparison the reference is to the subjects with diabetes diagnosed by OGTT alone. (B and C) Odds of diabetes being a leading (B) or contributing (C) cause of death stratified by diagnosis type. * indicates p < 0.011, ** indicates p < 0.001, and *** indicates p<2×10^−7^.

### Top features of interest in diabetes prediction

We first examined the 20 most important features from all the included variables, shown as a Shapley plots in Supplementary Figure 4. We found that the laboratory values that are affected by fasting (fasting plasma glucose, insulin, triglycerides, and iron) had a large predictive effect, ranking #1, #5, #8, and #16, respectively. Clinically-accessible vital signs included age, height, systolic blood pressure, waist circumference, diastolic blood pressure, and weight. The most important non-fasting labs were the A1c, urine albumin, percentage of segmented neutrophils, gamma-glutamyl transferase (GGT), creatinine, percentage of lymphocytes, lactate dehydrogenase (LDH), cholesterol, and alanine aminotransferase (ALT). The Poverty Ratio of the zip code of the participant was also in the top 20 variables (#19). Focusing on the more clinically applicable A1c+ model (Figure 2), which excluded the fasting labs fasting plasma glucose, insulin, triglycerides, and iron, and excluded the less-commonly obtained urine albumin, GGT, cholesterol, and LDH, we found that the aspartate aminotransferase (AST), hemoglobin, total protein, pulse, arm circumference, platelets, percentage of monocytes, and mean corpuscular volume were among the other features in the top 20 features of importance.

**Figure 2.**
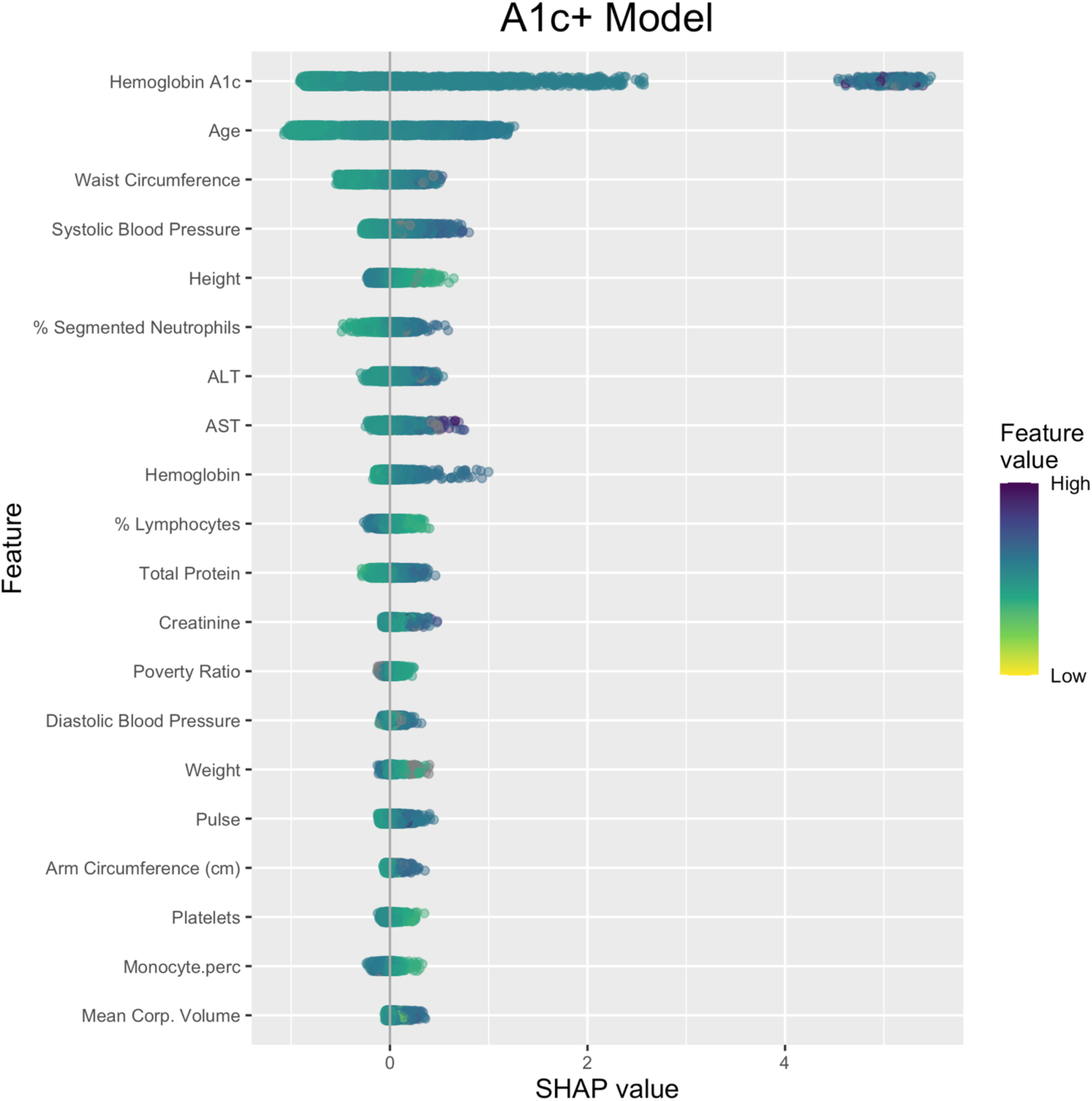
Plot of Shapley values for top 20 features of the A1c+ model. Each point represents a subject, with the Shapley (SHAP) value representing the contribution of a feature value to the difference between the actual prediction and the mean prediction without that feature. Positive values indicate a positive effect on the probability of diabetes, and negative values indicate a negative effect. RBC: Red Blood Cell; Corp: Corpuscular.

### Comparison of clinically accessible variables with A1c and FPG for diagnosing diabetes

We compared our non-fasting routine clinical feature model (A1c+) and A1c/FPG+ model (as described in *Methods)* to the current diagnostic thresholds of the A1c of 6.5% (A1c.Fixed) and FPG of 126 mg/dL (FPG.Fixed), as well as sensitivity analyses against models where the A1c and FPG were allowed to fluctuate to different thresholds (Figure 3, Supplementary Figure 5). Among the non-fasting approaches (A1c.Fixed, A1c, and A1c+), the A1c+ model outperforms the others, with an AUROC of 0.88 and sensitivity of 0.72 compared to the A1c AUROC of 0.83 and sensitivity of 0.66 and A1c.Fixed sensitivity of 0.37 (Figure 3A). Among the fasting approaches, the A1c/FPG+ model outperformed the simpler approaches, with an AUROC of 0.95 and a sensitivity of 0.87 compared to the FPG AUROC of 0.92 and sensitivity of 0.84, with a FPG.Fixed sensitivity of 0.48 (Figure 3B).

**Figure 3.**
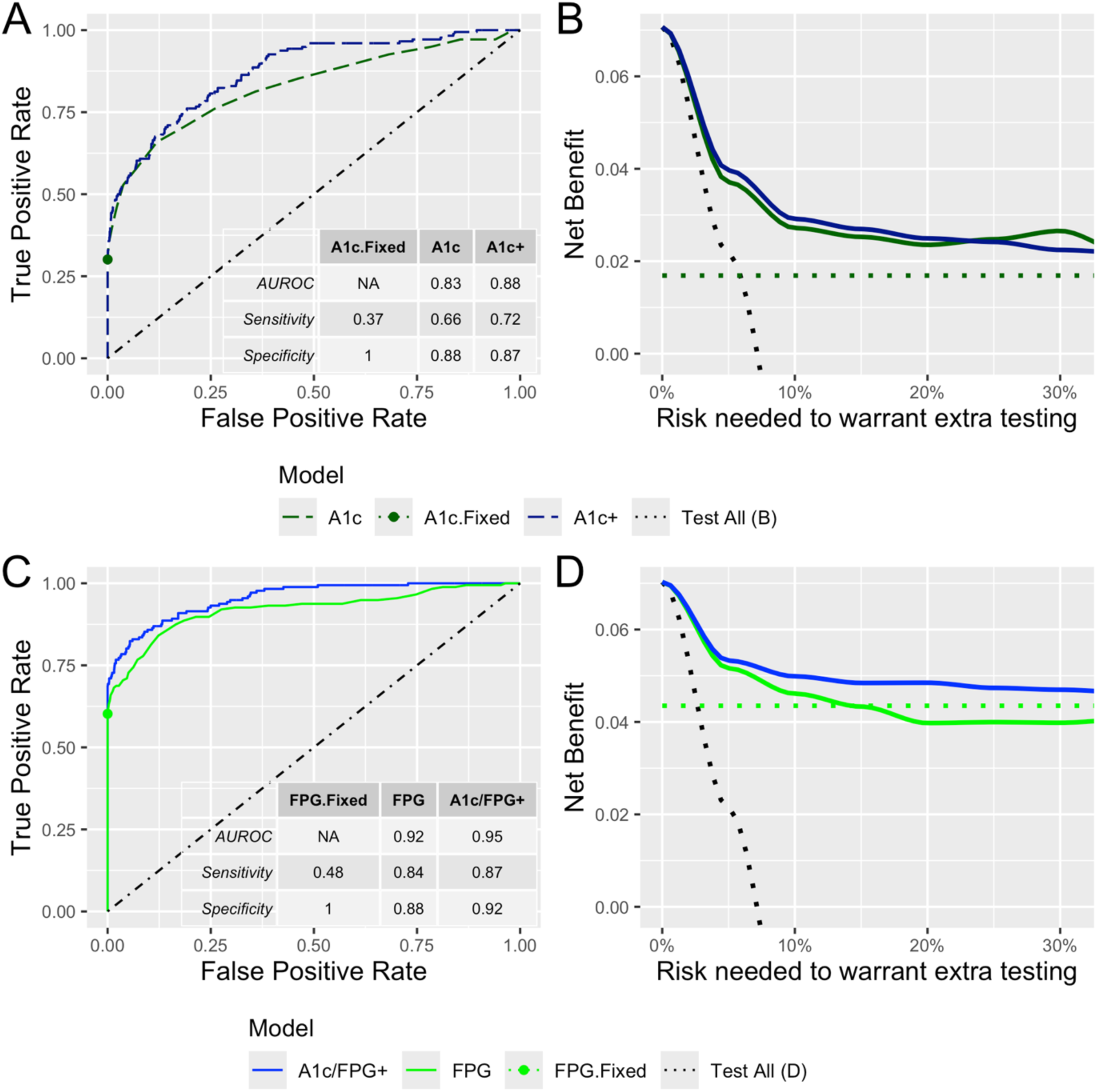
(A) Among non-fasting approaches, the routine clinical feature A1c+ model has higher sensitivity than the A1c with and without fixing the diagnostic threshold at 6.5% (A), while among the fasting models the A1c/FPG+ has higher sensitivity than the FPG with and without fixing the diagnostic threshold at 126 mg/dL (C). The A1c+ and A1c/FPG+ models also had superior AUROC (A and C). The Decision Curve Analysis shows that the A1c+ and A1c/FPG+ approaches outperform the A1c and FPG, respectively, from risk thresholds of 5-25%. Points in (A) and (C) indicate True Positive Rate and False Positive Rate of fixed thresholds for A1c=6.5% and FPG=126 mg/dL. Diagonal line in (A) and (C) indicates test characteristics of a random test. Dotted lines in (B) and (D) indicate the net benefit of testing all candidates or testing no candidates with oral glucose tolerance testing for diagnosis of diabetes.

To establish the utility of our models with a complementary method, we employed Decision Curve Analysis (Figure 3B). Decision Curve Analysis measures the net benefit difference between the approaches based on what risk of diabetes would be needed to convince patient or provider to order additional testing (in this case, an oral glucose tolerance test). For a diagnosis of diabetes, the benefit of the A1c+ model over the A1c and the benefit of the A1c/FPG+ model over the FPG extended from 5% to 25%, and across the entire range of risk compared to the A1c.Fixed.

For completeness, we included additional models, including a combined FPG+A1c XG Boost model and a model using all the available features (Full). The FPG+A1c model underperformed the A1c/FPG+ model by AUROC and sensitivity, and the Full model was equivalent to the simpler A1c/FPG+ model, justifying the use of the simpler A1c/FPG+ model (Supplementary Figure 5). We repeated our model building and analysis instead using the thresholds for pre-diabetes and diabetes together (A1c ≥ 5.7%, FPG ≥ 100 mg/dL, and 2hG ≥ 140 mg/dL). In that setting, the A1c+ model outperformed the A1c and the A1c/FPG+ performed equivalently to the FPG+A1c and Full model in outperforming the FPG (Supplementary Figure 6).

### Association of A1c/FPG+ model predictions with mortality

To assess the importance of our model predictions, we applied the A1c/FPG+ model to the 92,062 subjects in NHANES from 1999-2016 who did not have OGTT measurements (and therefore were not used for model training or evaluation), of whom 53,255 subjects had mortality data. We stratified the subjects into those with A1c- or FPG-diagnosed diabetes, those with sub-diagnostic A1c and FPG but with A1c/FPG+-predicted diabetes, and those with sub-diagnostic A1c and FPG but without A1c/FPG+-predicted diabetes (Figure 4A). We found that model-predicted diabetes (green line) was statistically indistinguishable from A1c- or FPG-diagnosed diabetes (red line) for their association with mortality when compared to subjects without diabetes or predicted diabetes (blue line), with HR = 8.2, p < 2*10^−16^ compared to HR = 8.2, p < 2*10^−16^, respectively (comparison HR=1.1, p<0.17). We found that prediction of diabetes among those without a diagnosis of diabetes was associated with higher odds of diabetes being a leading or contributing cause of death compared to those without a diagnosis of prediction of diabetes (0.89% vs. 0.25% and 6.63% vs 3.31%, respectively), though this was lower than the odds of diabetes as a leading or contributing cause of death for those with an A1c or FPG-based diagnosis of diabetes (Figure 4B & C).

**Figure 4.**
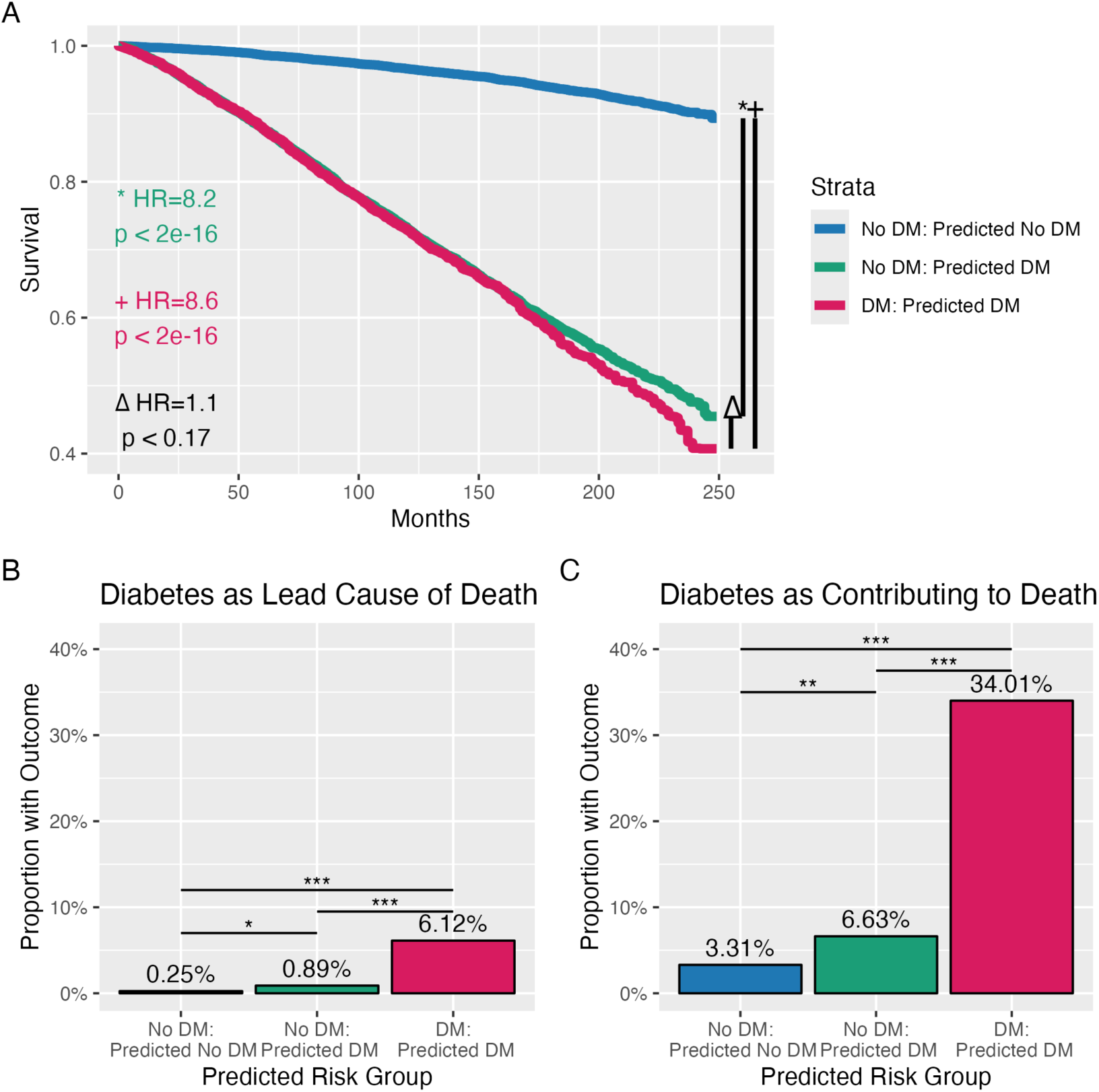
The subjects without diabetes at time of exam that were predicted to have diabetes via the A1c/FPG+ model had risk of mortality equivalent to those with diabetes (A), and a higher risk having diabetes as the leading cause (B) or contributing cause (C) of death compared to those not predicted to have diabetes by the A1cFPG+ model. Comparison of mortality (A), diabetes as a leading cause of death (B), and diabetes as a contributing cause of death (C) from NCHS data for NHANES 1999-2016 cohorts that did not have OGTT measurement stratified by subjects with a diagnosis of diabetes by A1c ≥ 6.5% or FPG ≥ 126 mg/dL (red) and subjects without a diagnosis of diabetes (A1c < 6.5% and FPG < 126 mg/dL) based on whether the A1c/FPG+ model predicts (green) or does not predict (blue) a diagnosis of diabetes. (A) Kaplan-Meier curves of mortality stratified by presence and prediction of diabetes. For the * and + comparisons the reference is to subjects with A1c < 6.5%, FPG< 126 mg/dL, and no prediction of diabetes. For the Δ comparison the reference is to the subjects with A1c < 6.5%, FPG< 126 mg/dL, and predicted diabetes. Prediction of diabetes in subjects without diabetes at time of exam increased risk of diabetes being the leading (B) or contributing (C) cause of death. * indicates p < 0.008, ** indicates p < 0.0003, and *** indicates p < 4×10^−10^.

## Discussion

In this study, we demonstrate that 1) OGTT-only diabetes has a higher (and equivalent if controlling for age, sex, and race/ethnicity) association with mortality compared to diabetes diagnosed by A1c or FPG, with equivalent risk of diabetes being the leading cause of death, 2) a machine learning model using features collected during standard medical visits can improve detection of diabetes, and 3) this machine learning-predicted diabetes associates with mortality equivalently to A1c- or FPG-diagnosed diabetes and associates with increased risk of diabetes being the leading or contributing cause of death compared those without predicted diabetes.

We have reaffirmed the importance of OGTT-only diabetes as having equivalent risk of death and diabetes-related death as diabetes diagnosed by A1c or FPG. Studies in earlier populations had established the predictive role of OGTT for mortality^26^, but this had not been examined in a modern 21^st^-century cohort. While the underlying pathophysiology of post-prandial hyperglycemia may differ from the elevated hepatic insulin resistance that drives increases in fasting plasma glucose, the risk of mortality appears to not change. From 2005-2016 over 1 in 100 individuals had OGTT-only diabetes. These individuals are missed by A1c and/or FPG screening approaches conventionally used, justifying the need for a means to identify these individuals that would otherwise be missed despite having equivalently bad outcomes.

We have developed a machine learning model that can easily integrate into clinical care, as it does not require additional vitals, demographics, or lab tests other than what are routine clinical features for a U.S. adult. Given this is information available in the electronic medical record, future integration of A1c/FPG+-type models into the electronic medical record (EMR) should guide screening. Additional models could be developed and validated that would tailor included clinical features to specific patient populations. This will be especially of use to general practitioners, who may not have the same familiarity with conditions that would require more extensive testing for diabetes with OGTT or bandwidth to pursue the work-up and diagnosis. In the setting of a sub-diagnostic A1c, an EMR model that calculated a high risk of diabetes could trigger an alert recommending fasting labs, and if the FPG was sub-diagnostic again an EMR model with a high risk could trigger a recommendation to obtain an OGTT, or at the very least repeat follow-up labs quickly (Supplementary Figure 7). While earlier detection and therefore earlier glucose control should lead to improved outcomes, further work and implementation will be needed to validate this.

The predictions of diabetes from the machine learning models are meaningful – subjects with the predictions with a diagnosis of diabetes have as high risk of mortality as those with diagnosed diabetes. There is strong evidence that the pathophysiological damage done by uncontrolled diabetes in the first few years after diagnosis has lifelong consequences. Diabetes was subsequently discovered in these subjects, as evidenced by diabetes appearing as a leading or contributing cause of death in the NCHS data. Earlier detection, especially now that there are multiple effective and complementary modalities for treating diabetes, could reduce this mortality. This paper and machine learning models offer a framework for providing that earlier detection.

These machine learning models and paper provide the groundwork for further work on this topic. This study externally validates the model using mortality data from NCHS – these machine learning models should be validated using other general population datasets as well as ideally chronic-disease specific datasets. Focusing on chronic diseases where the A1c is known to be inaccurate, such as a pregnancy, cystic fibrosis, pancreatitis, chronic liver disease, and chronic kidney disease, would fine-tune the models where there is a higher pre-test probability of OGTT-only diabetes. This would not necessarily require separate models: the underlying methodological framework of the decision-tree analysis should be able to fine-tune the branches of the tree in which creatinine (#12 in the A1c+ model), lipase, or ALT (#7 in the A1c+ model) is elevated, for example. Further work could also focus on the continuous aspect of prediction: here, we threshold on predicting diabetes vs. no diabetes, despite losing statistical power doing so^27^. Instead, a continuous risk score may better guide when to repeat standard testing as opposed to obtaining an OGTT, and the timeline in which to pursue these options.

## Conclusion

In this work, we have established that diabetes detected by OGTT but missed by A1c and FPG is carries equivalent risk of mortality. We have developed models that incorporate routine clinical features to detect diabetes that outperform the A1c and FPG, even if allowing recalibration of the A1c and FPG thresholds. We show the model-diagnosed diabetes in those without a diagnosis of diabetes have equivalent mortality to those with a diabetes diagnosis. Future work will need to validate, refine, and integrate these findings to improve our ability to personalize diabetes risk-stratification for patients.

## Data Availability

Data is available at https://github.com/alanlhutchison/NHANES-DMDx/tree/main

## Supplementary Methods

For National Health and Nutrition Examination Survey (NHANES) data between 1999 and 2004, we included the surveys *Demographics (DEMO), Blood Pressure (BPQ), Diabetes (DIQ)* and *Medical Conditions (MCQ) Questionnaires.* We additionally included *Body Measurements (BMX), Blood Pressure (BPX), Complete Blood Count (LAB25* for 1999-2000, *L25* for 2001-2004*), Biochemistry Profile (LAB18* for 1999-2000 and *L40* for 2001-2004*), Glycohemoglobin (LAB10* for 1999-2000 and *L10* for 2001-2004), and *Plasma Fasting Glucose, Serum C-peptide & Insulin (LAB10AM* for 1999-2000 and *L10AM* for 2001-2004) data. For 2001-2002 data, Alkaline Phosphatase measurements were taken from *L40_2_B*. We adjusted lab values to 2015-2016 measurements in accordance with NHANES recommendations based on changes in methodology and laboratory measurement techniques.

For NHANES data between 2005 and 2016, we included the surveys *Demographics (DEMO), Alcohol Use (ALQ), Blood Pressure (BPQ)*, *Diabetes (DIQ),* and *Medical Conditions (MCQ) Questionnaires.* We additionally included *Body Measurements (BMX), Blood Pressure (BPX)*, *Complete Blood Count (CBC), Biochemistry Profile (BIOPRO), Glycohemoglobin (GHB)*, *Magnesium and Related Tests (MGX),* Fasting Questionnaire (*FASTQX), Hepatitis A (HEPA, Hepatitis B (HEPB_S), Hepatitis C (HEPC),* and *Hepatitis E (HEPE)* data. We adjusted lab values to 2015-2016 measurements in accordance with NHANES recommendations based on changes in methodology and laboratory measurement techniques. Other than OGTT measurements, when generating the A1c+ and A1c/FPG+ models, we excluded features that were not present in both 1999-2004 and 2005-2016 cohorts.

XG Boost splits the data into separate branches at decision points (*e.g.* age greater than or less than 60 years), combines variables (*e.g.* left leg length and right leg length), and tunes thousands of parameters to optimize the model. Attempts to calibrate predictions using training (2005-2012), validation (2013-2014), and testing (2015-2016) datasets did not change the Brier scores used to evaluate quality of fit, so this calibration was not pursued, and training (2005-2014) and testing (2015-2016) data splitting was used as above. We used gradient boosted decision trees in *R* using the *xgboost* package and *caret* for hyperparameter tuning. The *AUC* and *pROC* packages were used for confusion matrix calculations, the *glm* package was used for binomial modeling, and the *dcurves* package was used for Decision Curve Analysis modeling. Plotting was done using *cowplot* and *ggplot2.* Kaplan-Meier curves were generated with the *survminer* package and Cox Proportional Hazard modeling was performed with the *survival* package. We employed survival analysis methods to compare mortality between groups in *R* using the *survey* and *survminer* packages. We compared odds of diabetes being a leading or contributing cause of mortality also using the *survey* package.

Incorporation of OGTT sample weights (WTSOG2YR), fasting weights (WTSAF2YR), examination weights (WTMEC2YR), and strata weights (SDMVSTRA, SDMVPSU) were employed for resampling and statistical testing. R Markdown files and data are available at the *GitHub* repository *alanlhutchison/NHANES-DMDx*, and the raw data is available from the Center for Disease Control and Prevention.

In total, the features included the following: basic information: vital signs collected in clinic, demographics, complete blood count, comprehensive metabolic panel, and A1c; fasting labs: fasting plasma glucose, fasting triglycerides, fasting insulin, and fasting iron; social determinants of health questionnaire; additional labs that may not be as frequently obtained in routine clinical care: the urine labs urine albumin and urine creatinine, the blood labs phosphorus, total cholesterol, gamma-glutamyl transferase (GGT), Hepatitis B core and surface antibody, globulin, lactate dehydrogenase, hydroxycontine, hepatitis A total antibody, and osmolality.

## Acknowledgements

We would like to thank Dr. Esra Tasali and Dr. Celeste Thomas for their feedback in preparation of this manuscript. No patients were involved in this study design or preparation of this manuscript. This study was evaluated by the University of Chicago IRB (study ID 25-1210) and deemed to be exempt from further review.

## Supplementary Figures and Tables

**Supplementary Table 1.**
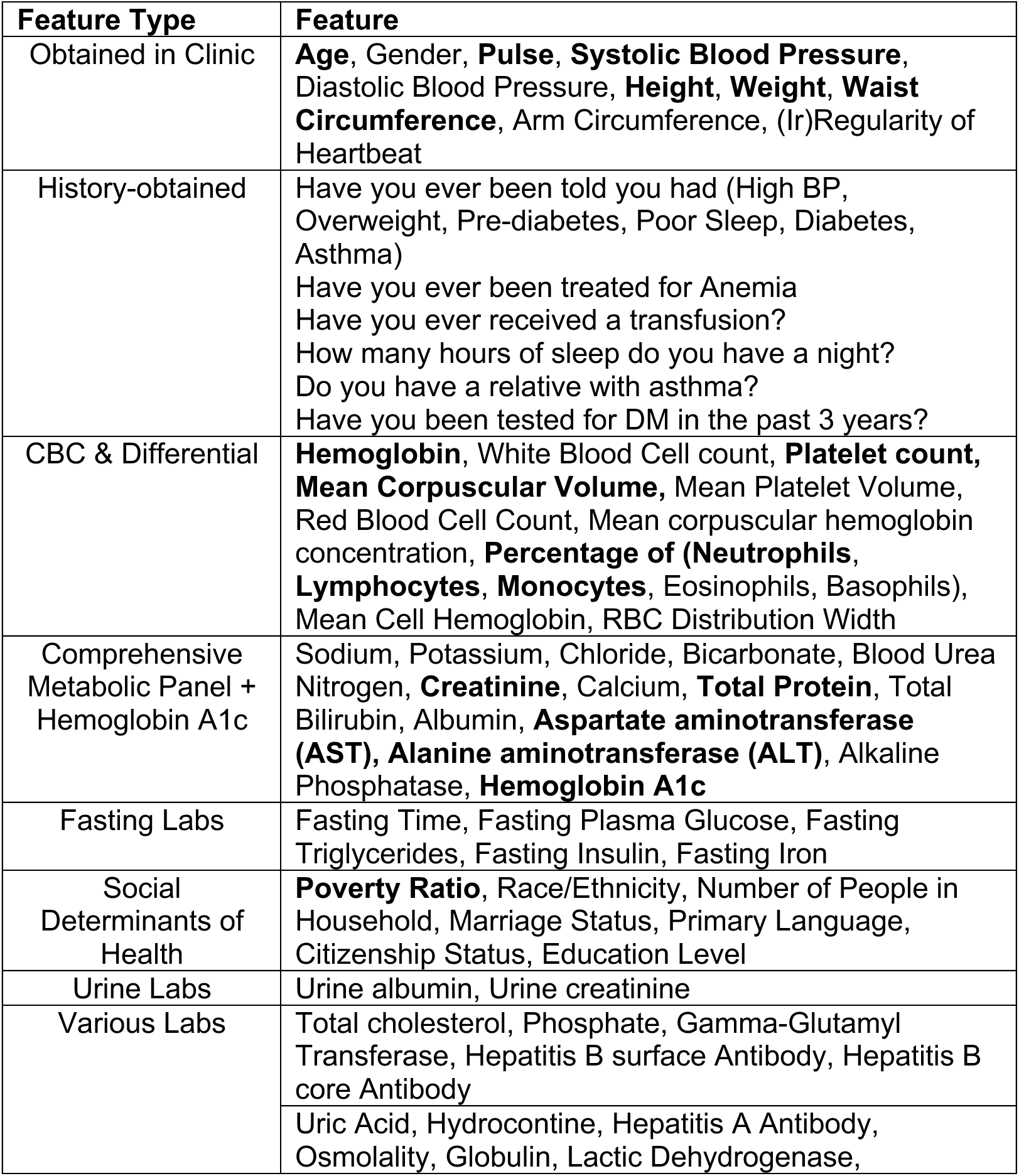
Features included in the machine learning Full model. Bolded are the top 20 features in the A1c+ model.

**Supplementary Table 2:** Redundant features excluded from analysis.

| Feature | Included Feature | Excluded Features |
| --- | --- | --- |
| Blood pressure | Systolic Blood Pressure average | BP site, BP arm, BP cuff peak pressure, BP enhancement usage |
| Length | Height | Arm length, Upper leg length |
| Weight | Weight | BMI |
| Language | Language | Interpreter usage, Proxy usage |
| Family Size | Number in family | Number in household |

**Supplementary Figure 1.**
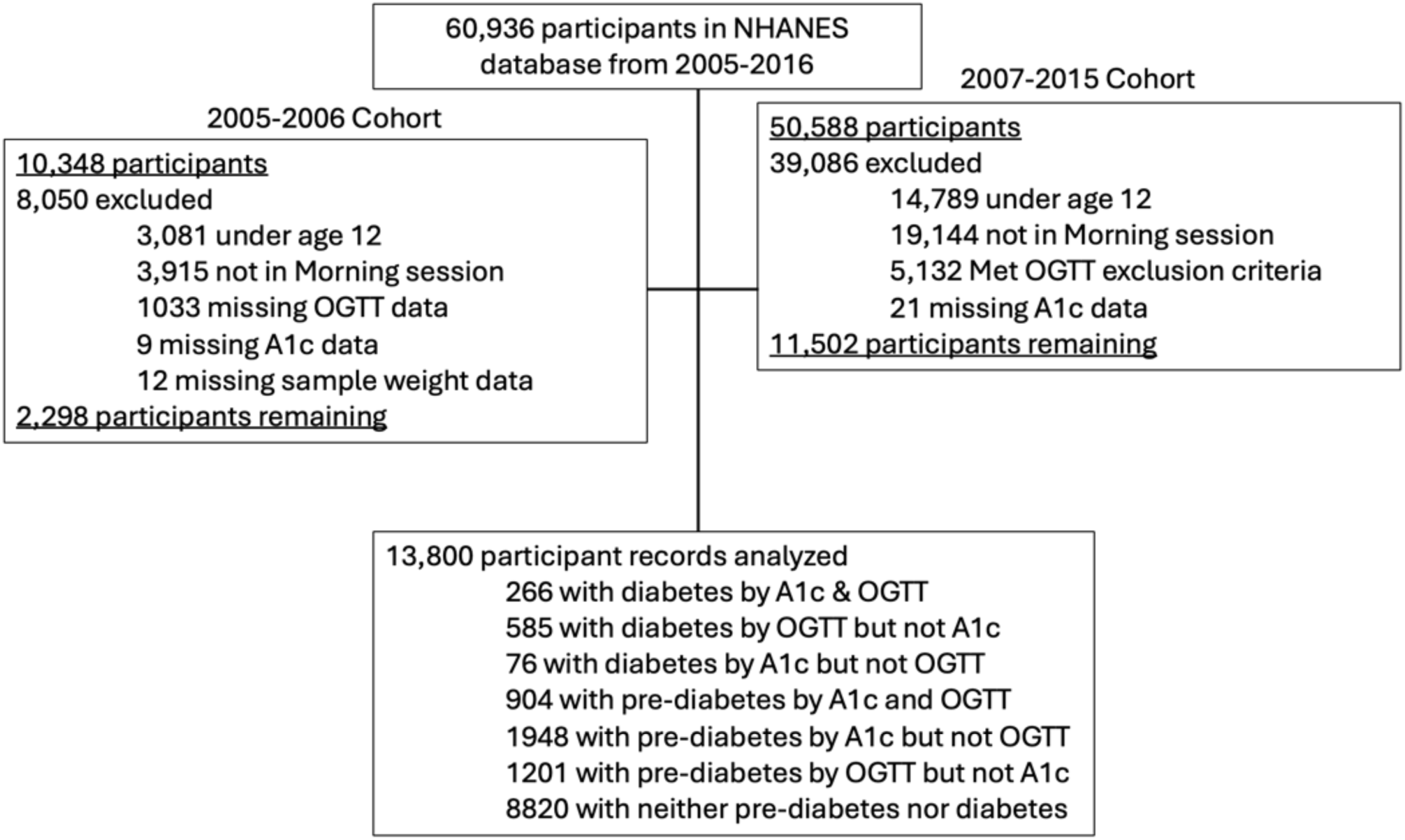
STROBE diagram.

**Supplementary Table 3.** Confusion matrix for the ability of the hemoglobin A1c, fasting plasma glucose, and two-hour oral glucose tolerance test thresholds to diagnose diabetes based on resampling 100,000 of the 13,800 subjects in the NHANES 2005-2016 dataset.

|  |  |  |
| --- | --- | --- |
| **A1c ≥ 6.5%** |  |  |
|  | **FPG ≥ 126 mg/dL** | **FPG < 126 mg/dL** |
| **2-hour Glucose ≥ 200** | 1.07% | 0.08% |
| **2-hour Glucose < 200** | 0.10% | 0.14% |
| **A1c < 6.5%** |  |  |
|  | **FPG ≥ 126 mg/dL** | **FPG < 126 mg/dL** |
| **2-hour Glucose ≥ 200** | 0.80% | 1.34% |
| **2-hour Glucose < 200** | 1.94% | 94.54% |

**Supplementary Figure 2.**
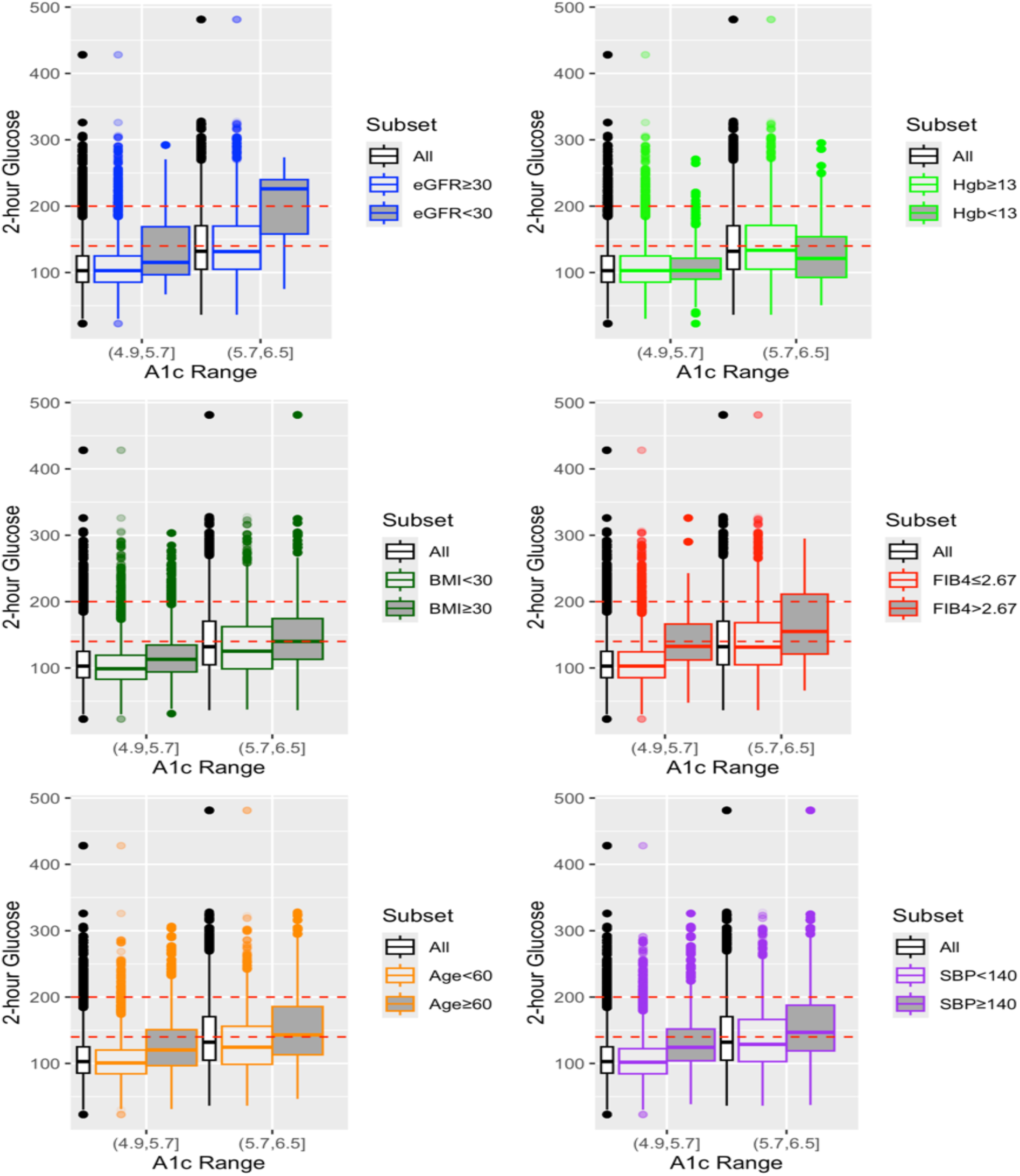
Relationship between Hemoglobin A1c and two-hour glucose stratified by A1c range. Red lines show thresholds used to diagnose pre-diabetes (140 mg/dL ≤2-hour glucose < 200) and diabetes (2-hour glucose ≥ 200). Data from NHANES 2005-2016 with hemoglobin A1c and two-hour glucose resampled with replacement 100,000 times based on OGTT sample weights. For Hgb comparison Hgb threshold of 13 g/dL was used for subjects classified as male and 11 g/dL for subjects classified as female. eGFR: Estimated Glomerular Filtration Rate (CKD-EPI), Hgb: Hemoglobin, BMI: Body Mass Index (in kg/m^2^), FIB4: non-invasive fibrosis risk^1^, SBP: Systolic Blood Pressure (in mmHg).

**Supplementary Figure 3.**
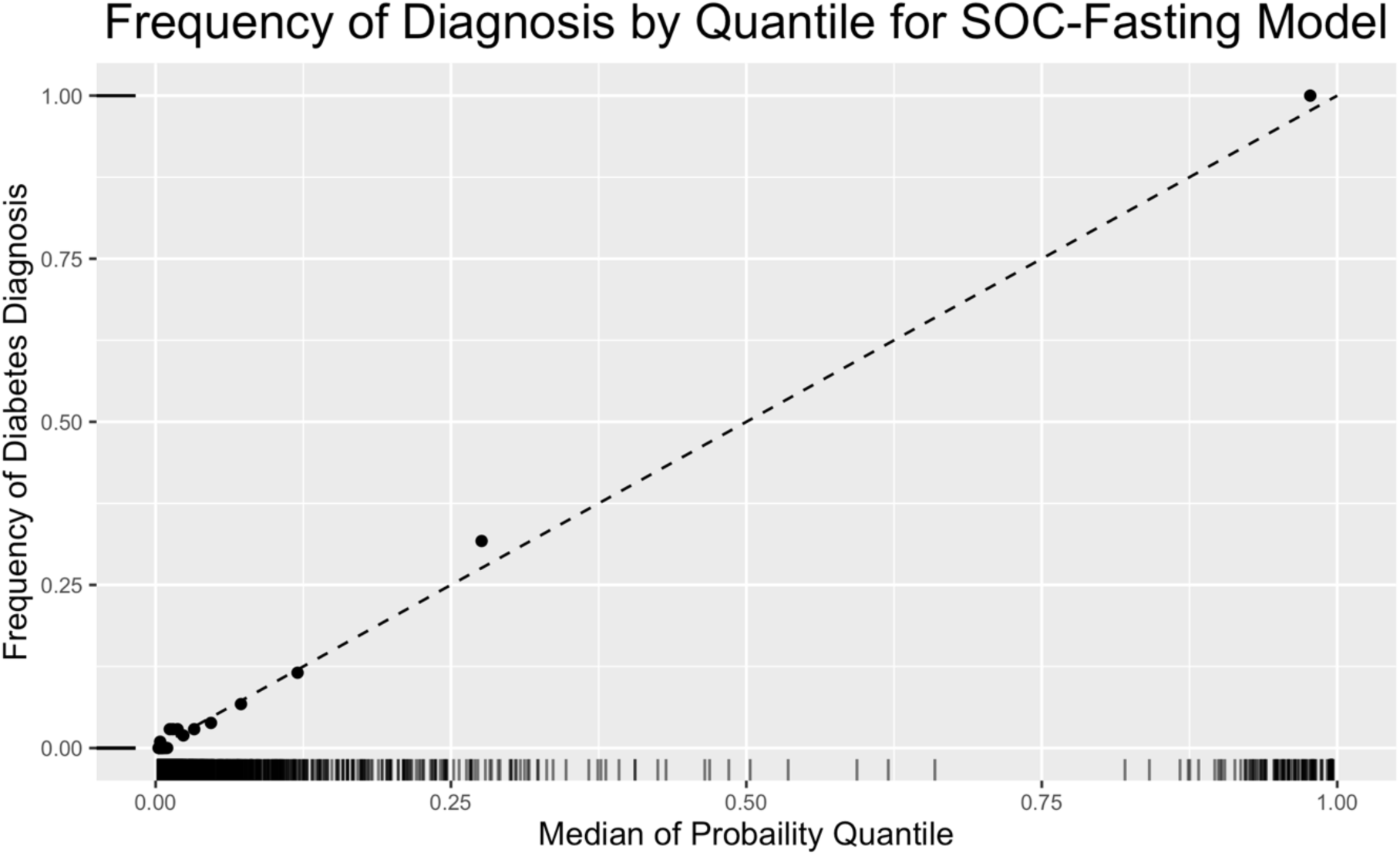
Calibration plot of Model-predicted diabetes, grouped by 10 quantiles (x-axis) compared to event rate within that group.

**Supplementary Figure 4:**
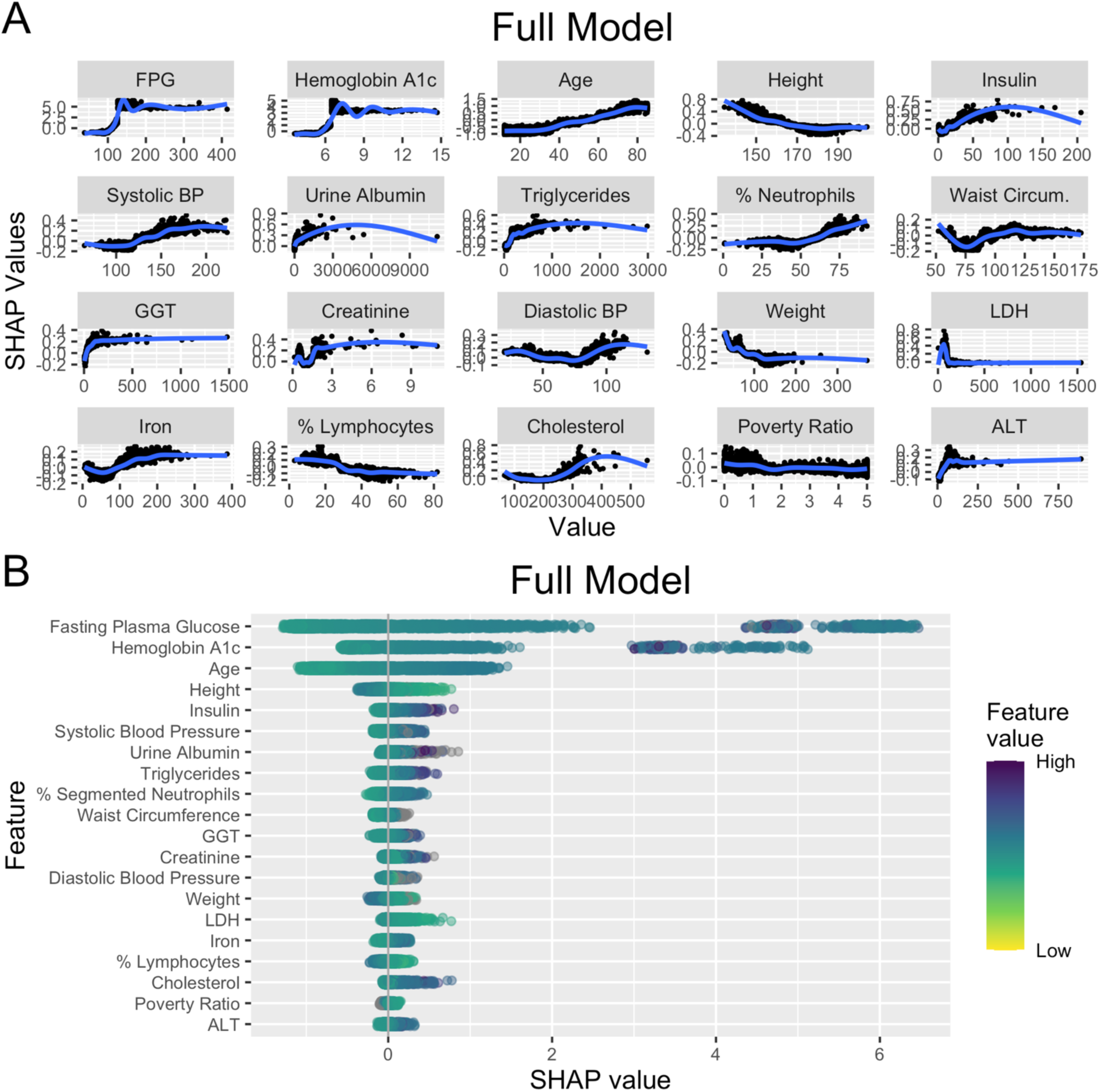
Plots of Shapley values for top 20 features of the Full model. Each point represents a subject, with the Shapley (SHAP) value representing the contribution of a feature value to the difference between the actual prediction and the mean prediction without that feature. Positive values indicate a positive effect on the predicted probability of diabetes, and negative values indicate a negative effect. Blue line indicates mean effect for a given value. FPG: Fasting Plasma Glucose; BP: Blood Pressure; GGT: Gamma-Glutamyl-Transferase; %. Neutrophils: Percentage Segmented Neutrophils; Mean Corp. Vol: Mean Corpuscular Volume; LDH: Lactic Dehydrogenase

**Supplementary Figure 5.**
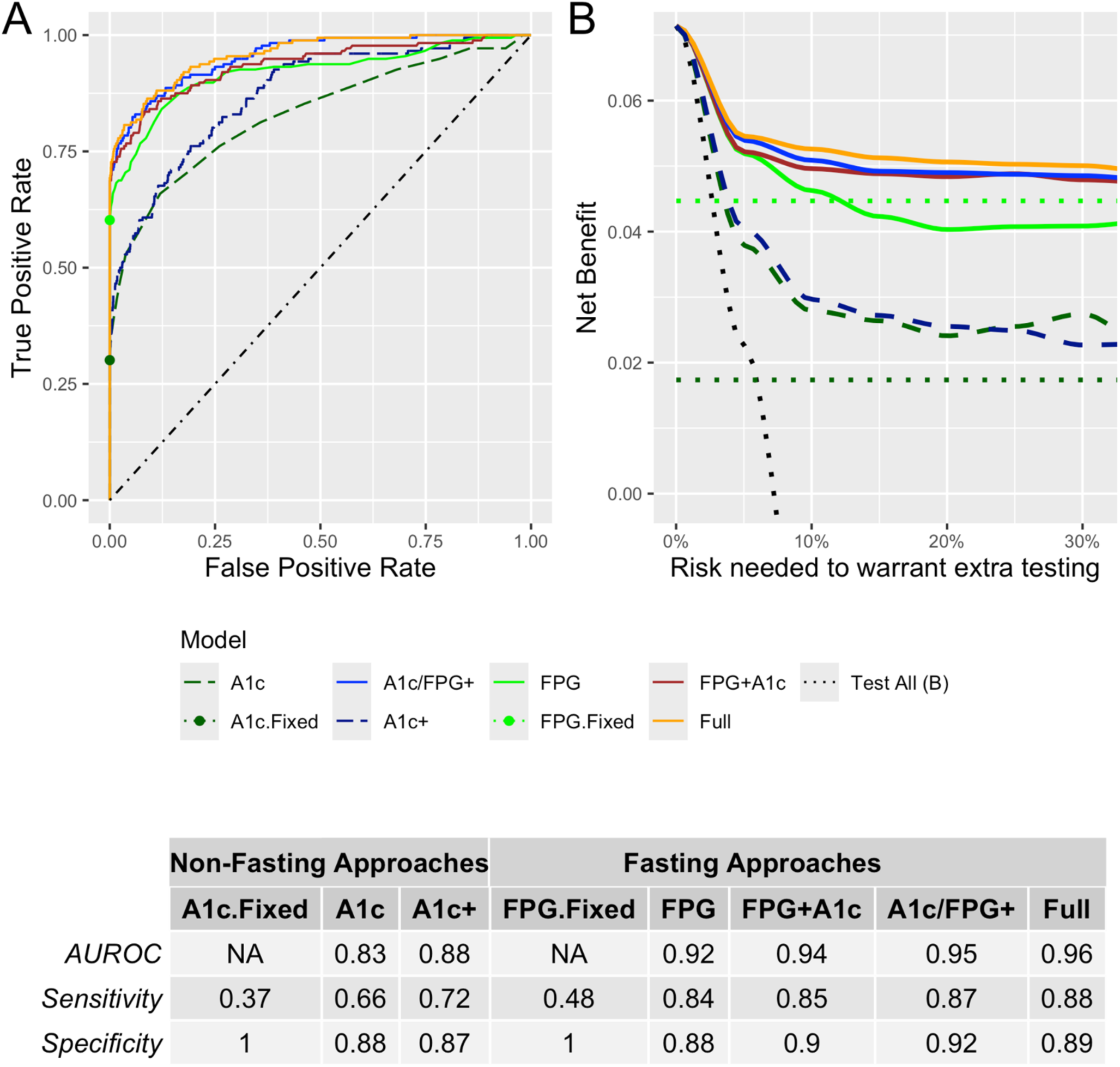
For diagnosis of diabetes, among non-fasting approaches, the non-fasting routine clinical feature A1c+ model has higher sensitivity than the A1c with and without fixing the diagnostic threshold at 6.5% (A), while among the fasting models the A1c/FPG+ model has higher sensitivity and AUROC than the FPG with and without fixing the diagnostic threshold at 126 mg/dL and the FPG+A1c model, and equivalent sensitivity and AUROC to the Full model. The Decision Curve Analysis (B) shows that the among non-fasting approaches the A1c+ is superior to the A1c, and among the fasting approaches the A1c/FPG+ and Full models are equivalently superior to the FPG and FPG+A1c models. Fasting approaches shown with solid lines, and non-fasting approaches shown with dashed lines. Points indicate True Positive Rate and False Positive Rate of fixed thresholds for A1c and FPG. Diagonal line in (A) indicates test characteristics of a random test. Dotted lines in (B) indicate the net benefit of testing all candidates (black) or testing no candidates (dark green: A1c, light green: FPG) with oral glucose tolerance testing for diagnosis of diabetes.

**Supplementary Figure 6.**
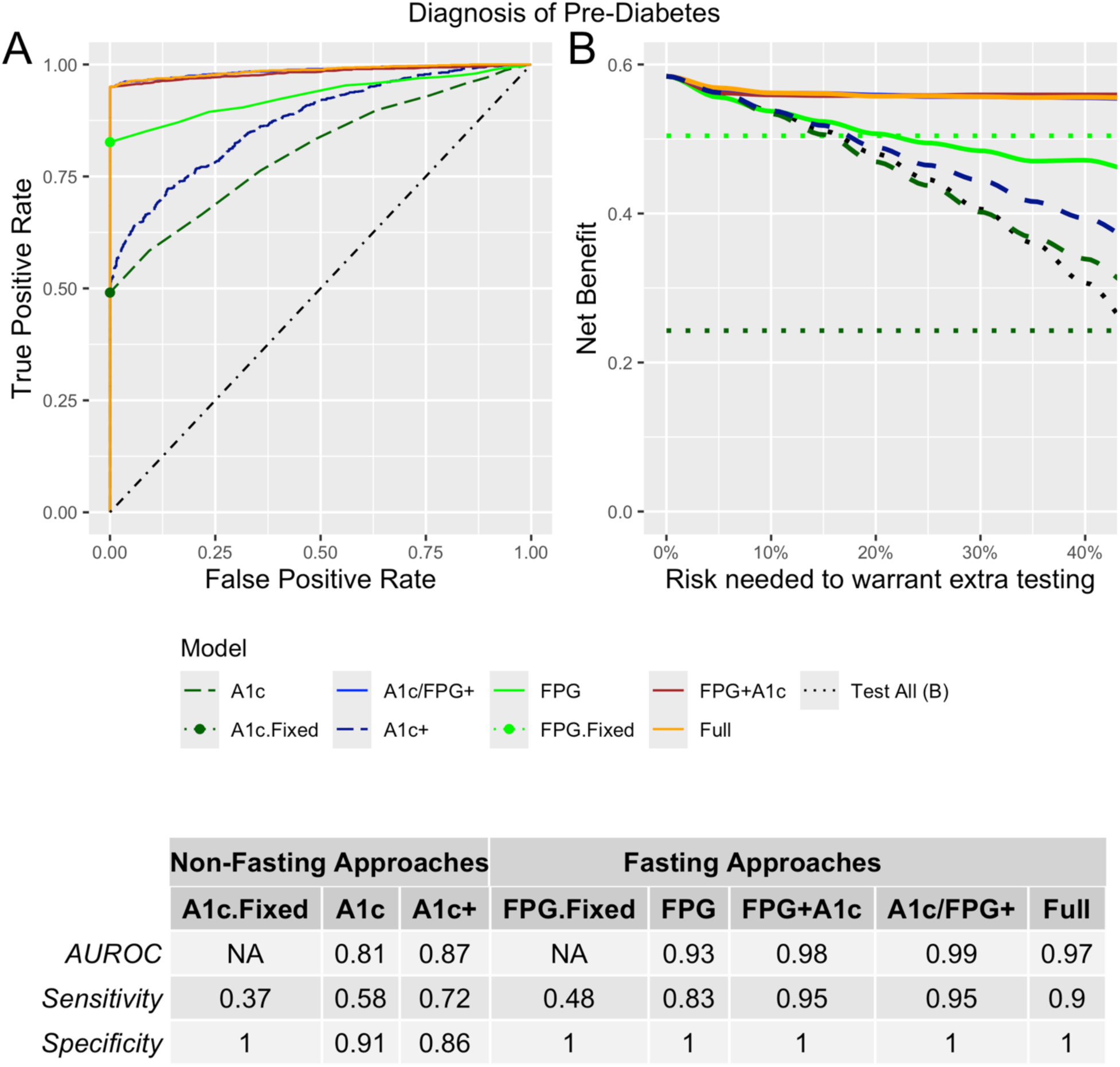
(A) Among non-fasting approaches, the non-fasting routine clinical feature model A1c+ had higher sensitivity than the A1c with and without fixing the diagnostic threshold at 6.5% (A, Table), while among the fasting models the A1c/FPG+ performed equivalently to the FPG+A1c and Full models and outperformed the FPG and FPG.Fixed models. The Decision Curve Analysis (B) likewise shows that the among non-fasting approaches the A1c+ is superior to the A1c and among the fasting models the A1c/FPG+ is equivalent to the FPG+A1c models and the FPG+A1c and Full models. Points indicate True Positive Rate and False Positive Rate of fixed thresholds for A1c and FPG. Diagonal line in (A) indicates test characteristics of a random test. Dotted lines in (B) indicate the net benefit of testing all candidates (black) or testing no candidates (dark green: A1c, light green: FPG) with oral glucose tolerance testing for diagnosis of pre-diabetes.

**Supplementary Figure 7.**
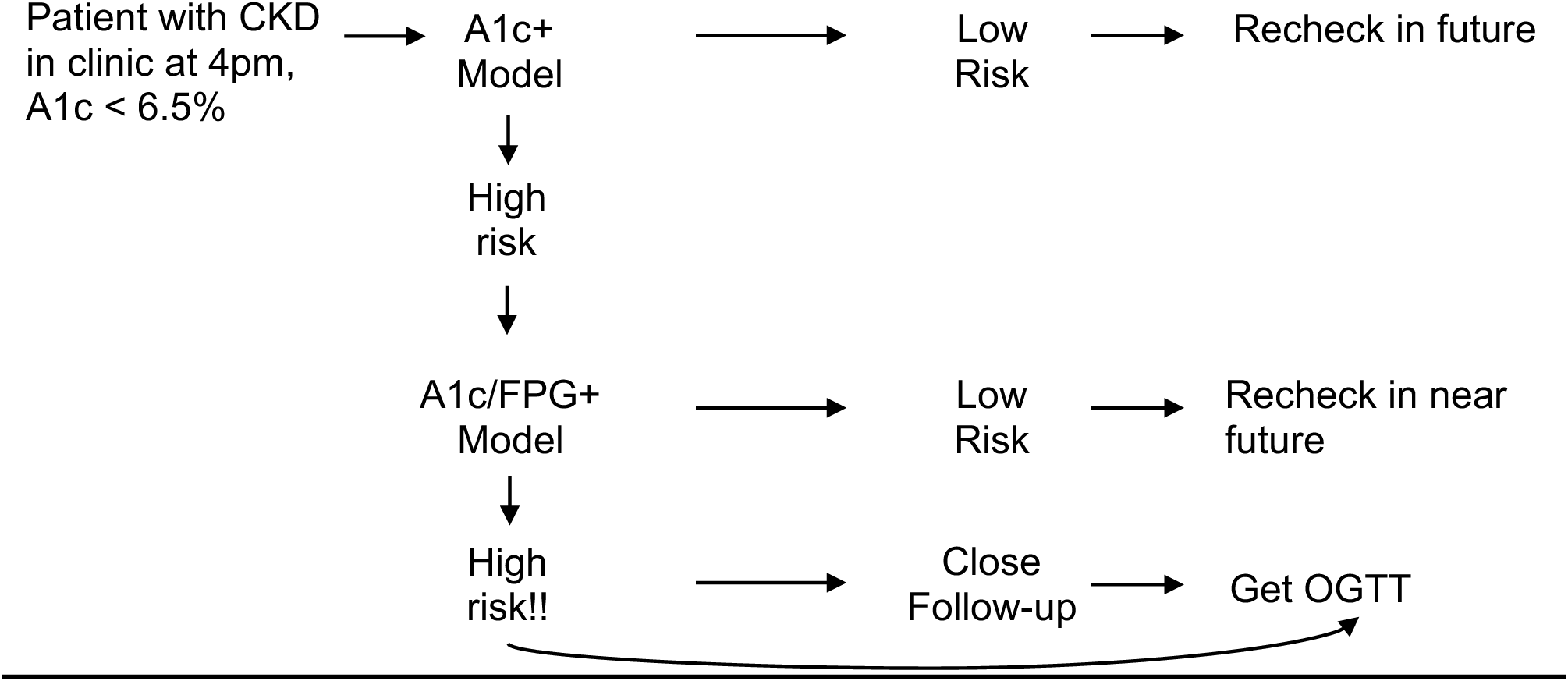
Schematic of conceptual framework of clinical application of A1c+ and A1c/FPG+ models.

